# Estimating long-term vaccine effectiveness against SARS-CoV-2 variants: a model-based approach

**DOI:** 10.1101/2023.01.03.23284131

**Authors:** Alexandra B Hogan, Patrick Doohan, Sean L Wu, Daniela Olivera Mesa, Jaspreet Toor, Oliver J Watson, Peter Winskill, Giovanni Charles, Gregory Barnsley, Eleanor M Riley, David S Khoury, Neil M Ferguson, Azra C Ghani

**Affiliations:** School of Population Health, Faculty of Medicine and Health, University of New South Wales, Sydney, NSW, Australia; MRC Centre for Global Infectious Disease Analysis, School of Public Health, Imperial College London, London, UK; Institute for Health Metrics and Evaluation, University of Washington, Seattle, USA; London School of Hygiene and Tropical Medicine, London, UK; Institute of Immunology and Infection Research, School of Biological Sciences, University of Edinburgh, UK; Kirby Institute, University of New South Wales, Sydney, NSW, Australia

## Abstract

With the ongoing evolution of the SARS-CoV-2 virus, variant-adapted vaccines are likely to be required. Given the challenges of conducting clinical trials against a background of widespread infection-induced immunity, updated vaccines are likely to be adopted based on immunogenicity data. We extended a modelling framework linking immunity levels and protection and fitted the model to vaccine effectiveness data from England for three vaccines (Oxford/AstraZeneca AZD1222, Pfizer-BioNTech BNT162b2, Moderna mRNA-1273) and two variants (Delta and Omicron) to predict longer-term effectiveness against mild disease, hospitalisation and death. We use these model fits to predict the effectiveness of the Moderna bivalent vaccine (mRNA1273.214) against the Omicron variant using immunogenicity data. Our results suggest sustained protection against hospitalisation and death from the Omicron variant over the first six months following boosting with the monovalent vaccines but a gradual waning to moderate protection after 1 year (median predicted vaccine effectiveness at 1 year in 65+ age group: AZD1222 38.9%, 95% CrI 31.8%-46.8%; BNT162b2 53.3%, 95% CrI 49.1%-56.9%; mRNA-1273 60.0%, 95% CrI 56.0%-63.6%). Furthermore, we predict almost complete loss of protection against mild disease over this period (mean predicted effectiveness at 1 year 7.8% for AZD1222, 13.2% for BNT162b2 and 16.7% for mRNA-1273). Switching to a second booster with the bivalent mRNA1273.214 vaccine against Omicron BA.1/2 is predicted to prevent nearly twice as many hospitalisations and deaths over a 1-year period compared to administering a second booster with the monovalent mRNA1273 vaccine. Ongoing production and administration of variant-specific vaccines are therefore likely to play an important role in protecting against severe outcomes from the ongoing circulation of SARS-CoV-2.

## Introduction

The rapid development and roll-out of SARS-CoV-2 vaccines has had a major effect on the health impacts of the global pandemic, substantially reducing COVID-19 cases, hospitalisations, and deaths^1–3^. Despite several vaccines showing high initial efficacy against infection with the Wuhan virus, the sequential emergence of variants of concern has substantially reduced the effectiveness of vaccines in blocking infection and onward transmission, although efficacy against severe outcomes has been more durable.^4–6^ The emergence and global spread of the Omicron variant and its subtypes has resulted in repeated infection due to waning and reduced effectiveness of vaccine- and infection-induced immunity.^7,8^ Omicron has now replaced prior variants globally and has been the dominant variant circulating for over 1 year, albeit with several emerging sub-variants.^9^ Two Omicron-specific bivalent vaccines, that include antigens representing both the original Wuhan virus and Omicron subtypes, are now available.^10^ These have demonstrated higher immunogenicity against the Omicron BA.1 subvariant and against the BA.4/BA.5 subvariants than the original vaccines.^11,12^

As SARS-CoV-2 continues to evolve, it is likely that both existing and updated variant-specific vaccines will lag viral antigenic evolution. Moreover, as is currently the case for influenza vaccines, decisions regarding investment in, or introduction of, new vaccines, as well as assessment of the need for further boosting, will likely be based on immunogenicity data rather than clinical trials. Obtaining reliable data on vaccine efficacy will be hampered by the high degree of infection-induced, broad-based antiviral immunity among most of the world’s population, making the identification of appropriate comparator groups challenging. This ongoing interaction between infection-induced immunity and vaccination (so-called “hybrid immunity”^13^) will also influence the effectiveness of vaccine booster programmes.

A method for estimating vaccine efficacy from immunogenicity data was proposed by Khoury *et al*.,^14^ who demonstrated that neutralizing antibody titres (NATs) could act as a correlate of protection across a range of SARS-CoV-2 vaccines. In this model, a non-linear dose-response model is estimated to relate NAT to protection against different clinical endpoints – capturing the more rapid decline in protection against mild disease that occurs as NAT declines over time compared to the slower decline in protection against more severe endpoints. Using this model, they subsequently predicted the loss of efficacy against emerging variants,^15^ as well as more recently the potential benefit of introducing variant-specific vaccines against a range of circulating variants.^16^ One of the limitations with a model based on NAT alone is that it does not capture the broader immune responses generated by vaccination (or infection) and how this may differ between vaccines.^17^ For example, studies have demonstrated that the adenovirus-vectored AstraZeneca AZD1222 vaccine induced broad T-cell responses even though the level of NAT induced following vaccination is lower than that of the mRNA vaccines.^18^ It is also suggested that inactivated whole virus vaccines (such as the VLA2001 vaccine manufactured by Valneva) should induce even more broadly based immune responses, which are in turn predicted to be both more durable and less susceptible to viral immune escape.^19^

Here we infer a simple model of immune waning and boosting directly from clinical endpoints. Using a similar model framework to that developed by Khoury *et al*.^14^ we infer the underlying immune dynamics by fitting to national-level vaccine effectiveness estimates from England. Although the immunological mechanisms of protection from severe disease are not entirely clear, NAT are a well-established mechanistic correlate of protection from infection. Whilst cellular immunity may play an additional role in protection from severe COVID-19, for simplicity we assume that the same immunological marker providing a surrogate for protection against infection can capture patterns of protection from severe disease. In so doing, we obtain estimates of vaccine efficacy against three endpoints – symptomatic mild disease, hospitalisation, and death – for combinations of three widely used vaccines. By incorporating follow-up through 2021 and 2022, we are able to estimate the longer-term duration of vaccine efficacy against both the Delta variant (dominant in 2021) and the Omicron BA.1/BA.2 variants (circulating in the first half of 2022).

## Methods

### Data

We used empirical estimates of vaccine effectiveness against mild disease (positive polymerase chain reaction (PCR) tests including symptomatic cases and asymptomatic infections detected through screening in schools and workplaces), hospitalisations (defined as admission recorded in the Emergency Care Dataset within 14 days of a positive test) and death (within 28 days of a positive test) with the Delta and Omicron BA.1/BA.2 variants from England.^20,21^ Data were available for three vaccines administered in England – the Oxford/AstraZeneca AZD1222 vaccine, the Pfizer-BioNTech BNT162b2 vaccine, and the Moderna mRNA-1273 vaccine – in various combinations. For our primary analysis we use data in the >65 age group from studies in which age-stratified estimates were provided.

We additionally extracted data on the immunogenicity (NAT) of the mRNA-1273.214 Moderna bivalent vaccine. These data are from an ongoing phase 2–3 study in which the vaccines were administered as a second booster dose in adults who had previously received three doses of the vaccine.^12^

### Immunological model

We followed the approach of Khoury *et al*.^14^ by considering the relationship between (here unobserved) immunity level (IL) over time, and protection against mild disease, hospitalisation and death.^14^ We first express an individual’s IL over time, *n*(*t*), as a biphasic exponential decay function where *n*_*ij*_ is the initial IL of vaccine *i* drawn from a log_10_-normal distribution at dose *j*. Based on B-cell dynamics, we assume an initial period of fast decay with half-life *h*_*s*_ (decay rate *π*1 = −*ln*(2)/*h*_*s*_) representing the combined biochemical decay of antibodies and the ongoing production of antibodies by circulating (mostly short-lived) plasma cells, followed by a second period of slow decay with half-life *h*_*l*_ (decay rate *π*_2_ = −*ln*(2)/*h*_*l*_), representing ongoing antibody production by long-lived plasma cells. This is represented by

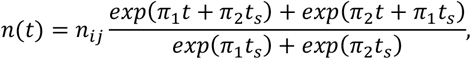

where *t* represents the time since the last dose, and *t*_*s*_ is the period of switching between the fast and slow declines. This results in a smoothed biphasic exponential waning of immunity levels. Whilst the original model is based on a simplification of B-cell dynamics, we note that bi-phasic patterns of immune decay also provide a good approximation to the more complex models of T-cell dynamics against other viruses in relation to longer-term protection following the initial acute infection.^22,23^

Following Khoury *et al*.^14^ we assume a logistic relationship between IL and vaccine effectiveness to capture time-varying vaccine protection against infection or mild disease (*m* = 1), hospitalisation (*m* = 2) and death (*m* = 3) given by the function

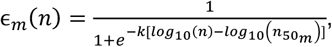

where ∈_*m*_ is vaccine effectiveness, *k* is the fitted shape parameter and 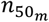 is the IL relative to convalescents required to provide 50% protection (from mild disease, hospitalisation, or death).^14^ Under this approach, we estimate different 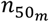 values for the different endpoints.

An alternative model is one in which protection against severe disease (hospitalisation and death) conditional upon infection or mild disease is assumed to be constant over time.^24^ We consider this model as a sensitivity analysis in the Supplementary Appendix.

We considered two different approaches to capture the effect of booster doses. In our main analysis, we consider a vaccine-dependent restoration of IL to a fixed dose-dependent level after each dose (including the two primary doses and one booster dose). Under this model, the restoration of protection is independent of the past decay in IL. Furthermore, the IL achieved at boost is independent of the vaccine regime used for the primary course, consistent with results from the COV-BOOST trial.^25^ As a sensitivity analysis, we assumed an alternative model in which there is a vaccine- and dose-dependent boost to IL, which therefore restores IL to a level that depends on both the magnitude of the boost and the level of IL achieved post dose 2. Under this model, the IL achieved at boost is therefore dependent on the vaccine regime used for the primary course but by being related to the level achieved post dose 2, does not change according to the time that has elapsed since dose 2. The results from this model are given in the Supplementary Appendix.

### Model fitting

Khoury *et al*.^14^ obtained parameters for this model of vaccine-induced protection by fitting the relationship between NAT data following dose 2 (which are based on the mean 28-day values reported in clinical trials relative to the mean titre for a convalescent individual) to clinical efficacy data from Phase III trials.^14^ We used these estimates as priors for our Bayesian model fitting.^13^ To fit the model jointly to estimates of vaccine effectiveness against the different variants (Delta and Omicron BA.1/2), we introduce a variant fold reduction (VFR) factor (to represent the degree of immune escape of Omicron) which scales the IL against Omicron compared with the baseline IL against Delta. This parameter is estimated in model fitting.

Our priors and their sources are summarised in **Table 1**. IL and VFR were transformed to the log_10_ scale, and all other parameters were fitted on a linear scale. We used Normal distribution priors on these scales with mean and standard deviations shown in **Table 1**. Model fitting was undertaken using parallel tempered MCMC methods using the DrJacoby R package with 400,000 samples.^26^

**Table 1:** Prior and posterior parameter estimates for the immunological model.

| Parameter | Symbol | Prior mean | Prior standard deviation | Posterior estimate for median (95% credible interval) | Reference for prior mean |
| --- | --- | --- | --- | --- | --- |
| **Immunity levels for each vaccine based on prior data on NAT** |  |  |  |  |  |
| **Oxford/AstraZeneca AZD1222 vaccine** |  |  |  |  |  |
| IL against Delta for dose 1 relative to convalescent | $n_{AZ,1}$ | $\log_{10}(0.5)$ | $\log_{10}(4)$ | 0.06 (0.04, 0.09) | - |
| IL against Delta for dose 2 relative to convalescent | $n_{AZ,2}$ | $\log_{10}((32/59)/3.9)$ | $\log_{10}(1.2)$ | 0.25 (0.19, 0.31) | 14,15,31 |
| IL against Delta for dose 3 relative to convalescent | $n_{AZ,3}$ | $\log_{10}(2)$ | $\log_{10}(4)$ | 0.45 (0.32, 0.61) | 25,32 |
| **Pfizer-BioNTech BNT162b2 vaccine** |  |  |  |  |  |
| IL against Delta for dose 1 relative to convalescent | $n_{PF,1}$ | $\log_{10}(0.5)$ | $\log_{10}(4)$ | 0.20 (0.15, 0.26) | - |
| IL against Delta for dose 2 relative to convalescent | $n_{PF,2}$ | $\log_{10}((223/94)/3.9)$ | $\log_{10}(1.2)$ | 0.52 (0.41, 0.63) | 14,15,33 |
| IL against Delta for dose 3 relative to convalescent | $n_{PF,3}$ | $\log_{10}(2)$ | $\log_{10}(4)$ | 0.72 (0.58, 0.91) | 25,32 |
| **Moderna mRNA-1273 vaccine** |  |  |  |  |  |
| IL against Delta for dose 1 relative to convalescent | $n_{MD,1}$ | $\log_{10}(0.5)$ | $\log_{10}(4)$ | 0.13 (0.09, 0.18) | - |
| IL against Delta for dose 2 relative to convalescent | $n_{MD,2}$ | $\log_{10}((654/158)/3.9)$ | $\log_{10}(1.2)$ | 0.73 (0.58, 0.91) | 14,34 |
| IL against Delta for dose 3 relative to convalescent | $n_{MD,3}$ | $\log_{10}(2)$ | $\log_{10}(4)$ | 0.91 (0.72, 1.16) | 25,32 |
| **Immune escape parameters** |  |  |  |  |  |
| Fold-reduction for Omicron relative to Delta (all vaccines) | VFR | 1.0 | $\log_{10}(4)$ | 6.5 (4.8, 8.9) | 35 |
| **Immunity level decay parameters** |  |  |  |  |  |
| Half-life of IL decay: short (days) | $h_s$ | 58 | 5 | 33 (29, 37) | 14,36 |
| Half-life of IL decay: long (days) | $h_l$ | 500 | 100 | 580 (415, 755) | 37 |
| Time period for switching (days) | $t_s$ | 90 | 20 | 86 (72, 97) | 14,36 |
| **Relationship between IL and protection** |  |  |  |  |  |
| IL relative to convalescent required to provide 50% protection from mild disease | $n_{50,1}$ | 0.33 | $\log_{10}(2)$ | 0.052 (0.034, 0.079) | 14 |
| IL relative to convalescent required to provide 50% protection from hospitalisation | $n_{50,2}$ | 0.05 | $\log_{10}(2)$ | 0.010 (0.005, 0.019) | 14 |
| IL relative to convalescent required to provide 50% protection from death | $n_{50,3}$ | 0.05 | $\log_{10}(2)$ | 0.010 (0.005, 0.020) | Prior based on hospitalisation<sup>14</sup> |
| Shape parameter | $k$ | 2.94 | 1 | 2.8 (2.4, 3.3) | 14 |

### Predicting bivalent vaccine effectiveness

As new COVID-19 vaccines will be evaluated based on immunogenicity data, we use NAT to provide a preliminary estimate of the effectiveness of the Moderna bivalent vaccine that is now in widespread use. To estimate the benefits for vaccine effectiveness, we compared the NAT against BA.1 for the Moderna mRNA.1273.214 bivalent vaccine to the equivalent NAT for the original mRNA.1273 vaccine.^12^ We used day 29 data against Omicron (BA.1) from those with no previous infection which gave a ratio of 2372/1473 = 1.61.^12^ This scaling of NAT was then applied (assuming it is representative of the scaling of the broader IL) to the fitted model to generate estimates of vaccine effectiveness against infection, hospitalisation, and death against BA.1/2.

## Results

**Figure 1.** shows the inferred relationship between immune level and protection against mild disease, hospitalisation and death for the Delta and Omicron variants. The shape of these curves is consistent with the observation that higher immune levels are required for protection against mild disease than for protection against the more severe endpoints (hospitalisation and death). This model generates a good fit to the observed vaccine effectiveness data for the 3 vaccines delivered in England and reproduces the differential rates of decline in vaccine effectiveness observed against both the Delta and Omicron variants over 1 year of follow-up (**Figures 2 and S1**). In sensitivity analyses, the alternative model in which protection against hospitalisation and death is conditional on protection against infection (as a constant scaling) provided the best overall fit to the data (**Table S2**). The relationship between immunity levels and protection against severe disease and death for this model differs from the main model, with less of a reduction in predicted vaccine effectiveness at lower immunity levels (**Figure S2**). Thus, this model predicts more sustained protection over time (**Figure S2**); however, this is inconsistent with more recent data which shows a decline in vaccine effectiveness that better aligns with our main model.^27^ The alternative boosting model produced a poorer fit to the data compared to our main model (**Table S2**) but generated a similar relationship between immunity levels and protection (**Figure S3**).

**Figure 1:**
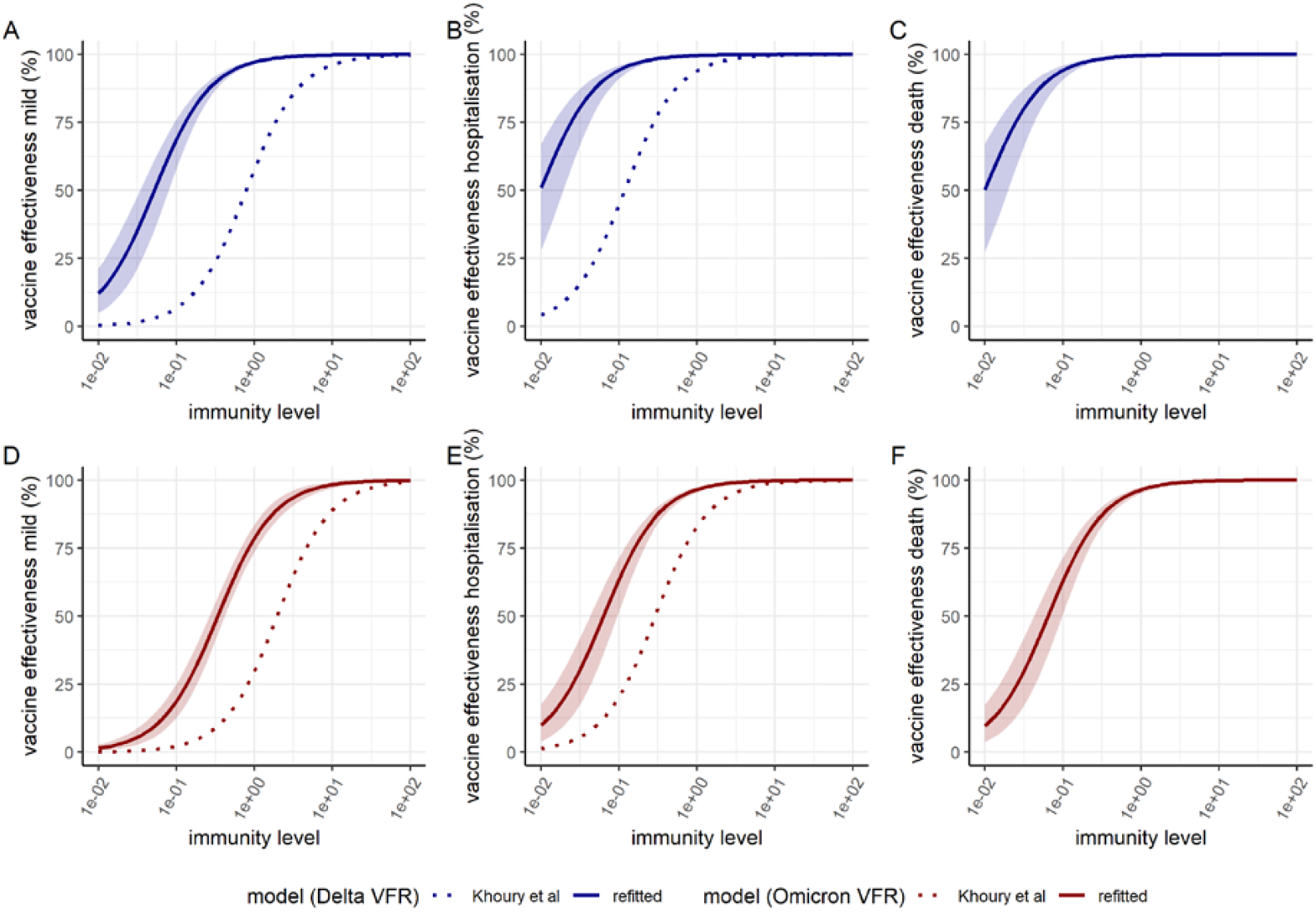
Dose-response curves estimated from fitting to vaccine effectiveness data for the relationship between immunity level (IL, x-axis) and vaccine effectiveness against mild disease (A, D), hospitalisation (B, E) and death (C, F). Panels A–C show vaccine effectiveness against the Delta variant whilst panels D–F show vaccine effectiveness against the Omicron/BA.1 variant. The solid lines show the posterior median estimates and colour bands the 95% credible interval. The dotted lines show the dose-response curves using the original efficacy model presented in Khoury *et al*.^14^ adjusted for the Delta and Omicron variants respectively by using the variant fold reductions (VFRs) reported in Cromer *et al*. and Khoury *et al*.^15,28^

**Figure 2:**
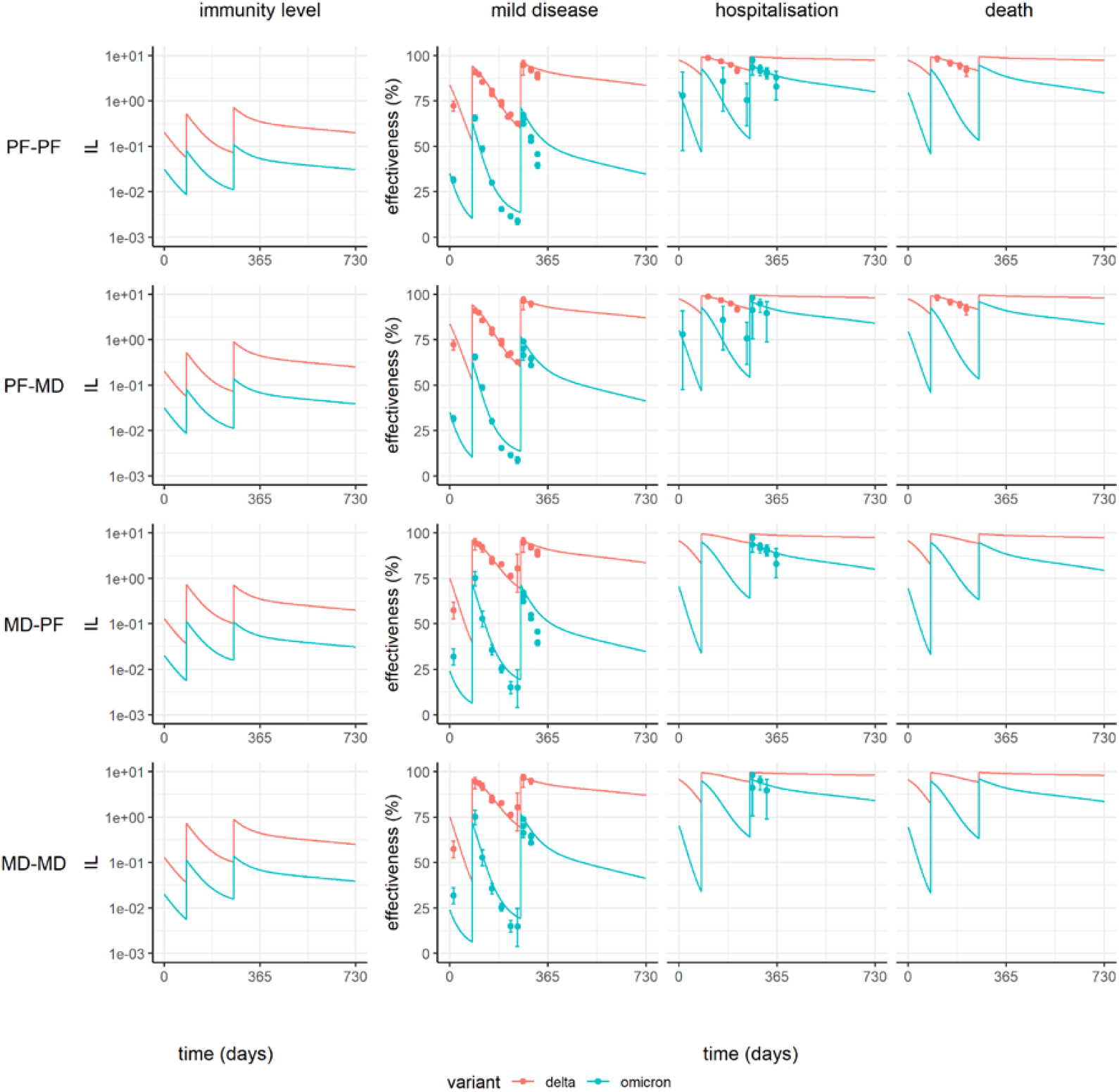
Predicted vaccine effectiveness over time for combinations of schedules for the original Pfizer-BioNTech BNT162b2 (PF) and Moderna mRNA-1273 (MD) vaccines. Plots show neutralizing immunity level (IL) in the left column, alongside efficacy against mild disease, hospitalisation, and death on the right. Neutralization and protection against the Delta and Omicron variants are shown in red and blue respectively. Four regimens are shown: PF delivered for three doses (PF-PF); two doses of PF and a booster dose of MD (PF-MD); two doses of MD and a booster dose of PF (MD-PF); and MD delivered for three doses (MD-MD). The solid lines show the posterior median fitted model estimate, and the points show estimates of vaccine effectiveness against three endpoints using data from England.^20,38^

We estimate a 6.5-fold (95% CrI 4.8–8.9) reduction in IL (induced by vaccination with the original Wuhan strain of the virus) against the Omicron variant relative to the Delta variant. The dashed lines on **Figure 1** show the expected relationship using the Khoury *et al*.^14^ model for the Delta and Omicron viruses respectively, using meta-analyses of NAT (applying a 3.9-fold reduction from the Wuhan virus to Delta^15^ and 9.7-fold reduction from the Wuhan virus to the Omicron BA.1 variant^28^, respectively). For both the Delta and the Omicron variants, applying the fold reductions estimated from immunogenicity data to the relationship inferred against the Wuhan virus results in a more pessimistic prediction of vaccine effectiveness than was inferred from fitting to the clinical data.

The fitted model parameters are summarised in **Table 1**. In terms of comparative effectiveness of the three vaccines, we estimate a trend with the highest IL generated with mRNA-1273 followed by BNT162b2and then AZD1222, consistent with the empirical data which shows higher levels of peak protection following both dose 2 and dose 3. The half-life of IL during the initial more rapid period of decay is estimated to be 33 days (95% Credible Interval (CrI) 29–37 days), shorter than the estimate of 58 days estimated by Khoury *et al*.^14^ for neutralization titre decay following infection although our model structure includes a more gradual transition to the slow delay over an 86-day period. The estimate of the half-life for the subsequent longer period of decline of 580 days (95% CrI 415–755 days) was consistent with the 500 days assumed by Khoury *et al*.,^14^ and with the wide uncertainty bounds indicating a remaining degree of uncertainty in the longer-term durability of protection.

**Table 2** shows estimates of vaccine effectiveness over time against the Omicron variant (comparative estimates against the Delta variant are given in **Table S2**). We estimate that 180 days after boosting, effectiveness against hospitalisation with Omicron declines to 49.5% (95% CrI 42.7%–56.7%) for AZD1222, 69.8% (95% CrI 68.0%–71.9%) for mRNA-1273 and 63.7% (95% CrI 61.7%–65.7%) for BNT162b2, consistent with estimates from 105+ days of follow-up in the UK.^29^ One year after vaccination, these levels are predicted to decline further, resulting in relatively low protection against infection or mild disease, and moderate protection against hospitalisation (38.9%, 95% CrI 31.8%–46.8% for AZD1222; 60.0%, 95% CrI 56.0%–63.6% for mRNA-1273; 53.3%, 95% CrI 49.1%–56.9% for BNT162b2).

**Table 2:** Estimated vaccine effectiveness against mild disease, hospitalisation and death for Oxford/AstraZeneca AZD1222, Pfizer-BioNTech BNT162b2 and Moderna mRNA-1273 vaccine regimens as a function of time since dose 2 or booster. Estimates are shown for the Omicron variant with the original vaccines; estimates for the Delta variant are shown in **Table S2**. Values shown are the posterior median and 95% credible intervals.

| Vaccine | Days post dose 2 |  | Days post booster |  |  |  |  |  |  |
| --- | --- | --- | --- | --- | --- | --- | --- | --- | --- |
|  | 90 | 180 | 30 | 60 | 90 | 120 | 150 | 180 | 365 |
| *Mild disease* |  |  |  |  |  |  |  |  |  |
| Oxford/AstraZeneca AZD1222 | 12 (11.5-12.4) | 6.1 (5.8-6.4) | 42.3 (35.7-49.3) | 29.9 (24.4-36) | 21.5 (17.2-26.6) | 16.4 (12.9-20.6) | 13.4 (10.4-16.9) | 11.6 (8.9-14.8) | 7.8 (5.8-10.3) |
| Moderna mRNA-1273 | 33.1 (32.2-34.2) | 19.1 (18.2-20.2) | 63.4 (62.7-64.4) | 50.1 (49.2-51.4) | 39.2 (38.2-40.6) | 31.6 (30.6-33) | 26.7 (25.7-28) | 23.6 (22.5-24.9) | 16.7 (14.6-18.7) |
| Pfizer-BioNTech BNT162b2 | 24.8 (24.1-25.3) | 13.6 (13-14.1) | 56.7 (56.2-57.5) | 43.3 (42.5-44.1) | 32.9 (32.1-33.8) | 26 (25.1-26.8) | 21.6 (20.9-22.5) | 19 (18.1-20) | 13.2 (11.4-14.8) |
| *Hospitalisation* |  |  |  |  |  |  |  |  |  |
| Oxford/AstraZeneca AZD1222 | 50.6 (48.9-52.1) | 32.8 (31-34.4) | 84.6 (80.7-87.9) | 76.1 (70.8-80.8) | 67.2 (60.9-73.1) | 59.5 (52.8-66.1) | 53.6 (46.8-60.6) | 49.5 (42.7-56.7) | 38.9 (31.8-46.8) |
| Moderna mRNA-1273 | 78.7 (77.6-80.1) | 63.9 (61.9-65.9) | 92.8 (92.4-93.3) | 88.3 (87.6-89) | 82.9 (81.9-84) | 77.6 (76.3-79) | 73.1 (71.6-75) | 69.8 (68-71.9) | 60 (56-63.6) |
| Pfizer-BioNTech BNT162b2 | 71.1 (69.7-72.4) | 54 (52-55.9) | 90.8 (90.3-91.3) | 85.1 (84.4-85.9) | 78.6 (77.6-79.8) | 72.4 (71.1-73.8) | 67.4 (65.8-69.1) | 63.7 (61.7-65.7) | 53.3 (49.1-56.9) |
| *Death* |  |  |  |  |  |  |  |  |  |
| Oxford/AstraZeneca AZD1222 | 49.7 (46-53.6) | 32 (28.8-35.6) | 84.1 (79.8-87.8) | 75.5 (69.7-80.7) | 66.4 (59.5-72.9) | 58.6 (51.3-65.7) | 52.7 (45.3-60.3) | 48.6 (41.3-56.4) | 38.1 (30.5-46.4) |
| Moderna mRNA-1273 | 78.1 (75.6-80.9) | 63 (59.6-66.9) | 92.6 (91.6-93.7) | 87.9 (86.4-89.6) | 82.3 (80.3-84.8) | 76.9 (74.3-80) | 72.4 (69.5-75.9) | 69 (65.8-72.8) | 59.2 (54-64) |
| Pfizer-BioNTech BNT162b2 | 70.4 (67.3-73.6) | 53.1 (49.4-57.2) | 90.5 (89.2-91.8) | 84.6 (82.7-86.7) | 78 (75.5-80.8) | 71.7 (68.7-75) | 66.6 (63.4-70.3) | 62.8 (59.2-66.8) | 52.4 (47.2-57.3) |

Using the immunogenicity data from those with no prior infection in Chalikas *et al*.,^12^ we calculate a 1.61-fold increase in NAT against BA.1 after boosting with mRNA.1273.214 versus mRNA.1273. The resulting estimates of vaccine effectiveness against BA.1 after applying this increased level of immunogenicity to the ILs in our model are shown in **Table 3** with the estimated vaccine effectiveness curves following a 4^th^ booster dose shown in **Figure 3A**. Compared with boosting with the original mRNA.1273 vaccine, we estimate relatively little difference in levels of protection against hospitalisation and death shortly after the boost between the two vaccines, relative to the much more substantial impact that administering any booster dose has compared to not boosting (**Figure 3B**). However, this pattern is not sustained over time with a predicted more rapid drop in efficacy against both mild disease and hospitalisation endpoints during 1 year of follow-up with the original vaccine compared to the bivalent vaccine, such that efficacy against hospitalisation reduces to 60.0% (95% CrI 56.0%–63.6%) 365 days following the fourth dose with the original vaccine but remains substantially higher (77.9%, 95% CrI 75.0%–80.4%) with the bivalent vaccine. Across a 1-year period, the overall impact is such that just over half of the additional protection is predicted to occur from delivering a 4^th^ booster dose compared to no booster, and the other half from switching to the bivalent vaccine rather than continuing with the original vaccine (**Figure 3B**).

**Table 3:**
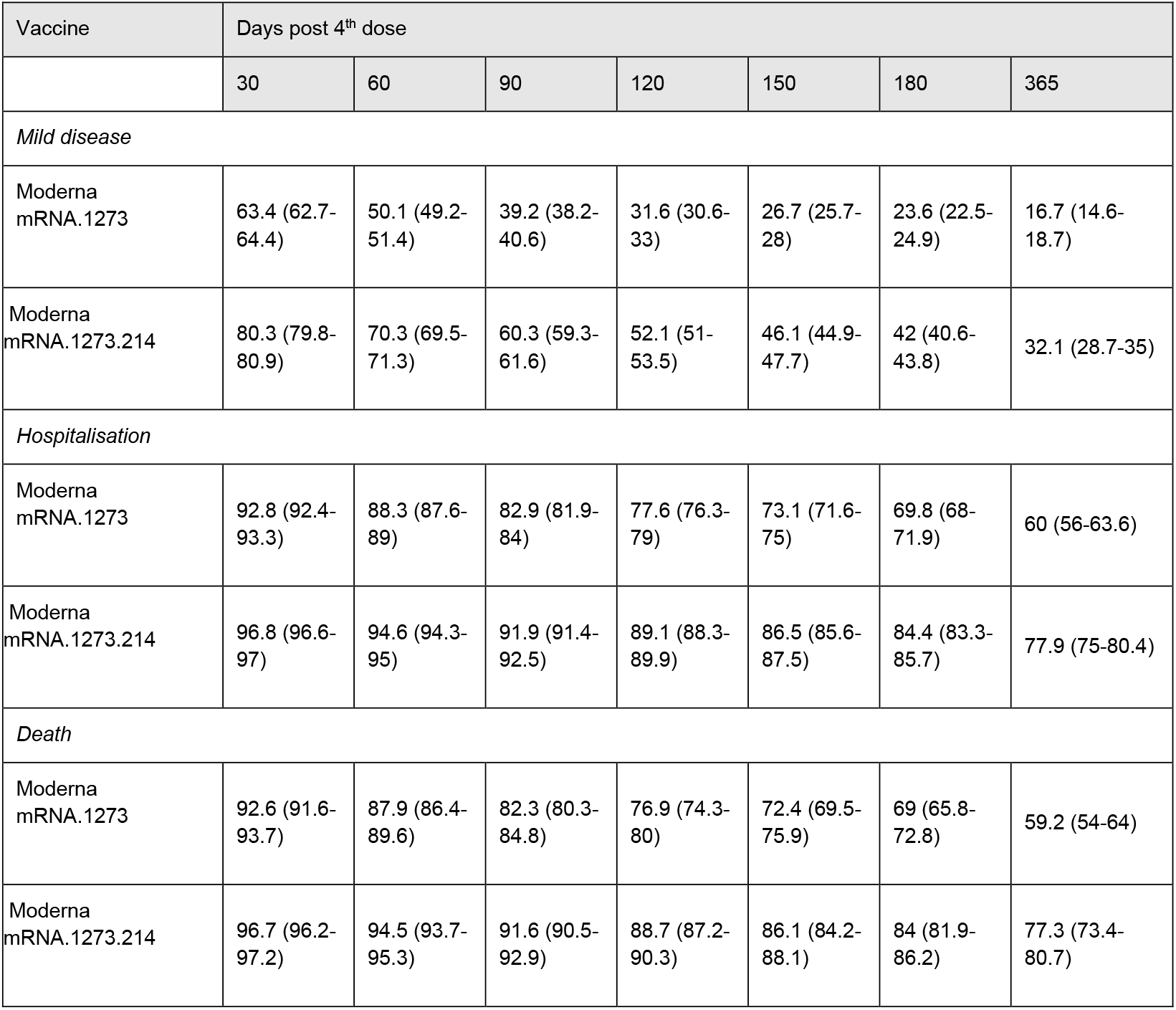
Estimated vaccine effectiveness against mild disease, hospitalisation and death from BA.1 for the original (mRNA.1273) and bivalent (mRNA.1273.214) Moderna vaccines as a function of time since a fourth dose. Values shown are the posterior median and 95% credible intervals.

**Figure 3:**
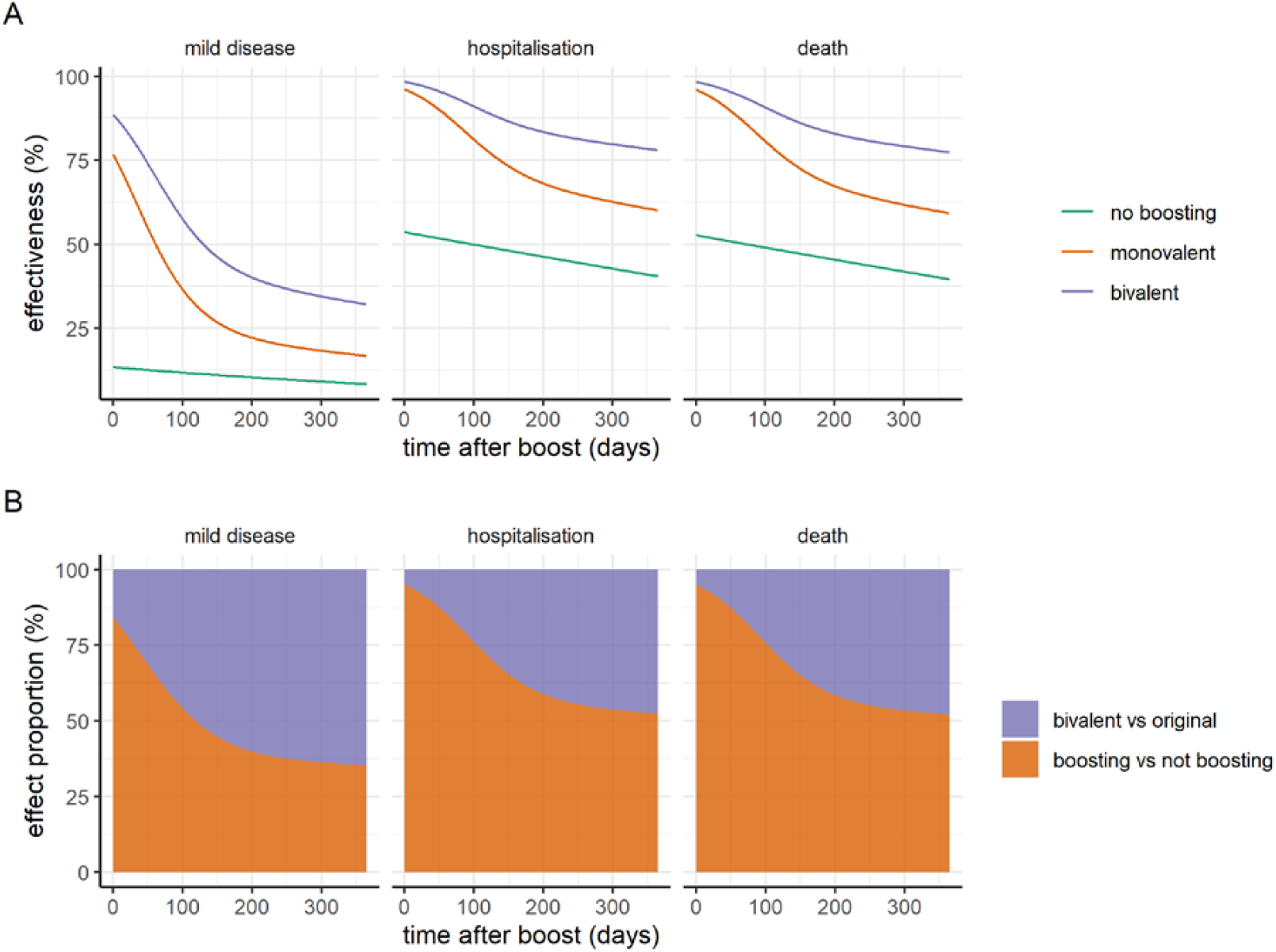
Model predicted vaccine effectiveness over time following a fourth dose boosting with either the monovalent or bivalent Moderna vaccine (mRNA.1273 or mRNA.1273.214 respectively), compared to no further boosting after the third dose. (A) Vaccine effectiveness against mild disease, hospitalisation, and death, for three doses only (green line), a fourth dose with the current Moderna vaccine (orange line), and a fourth dose with the bivalent Moderna vaccine (purple line). Estimates are against the Omicron BA.1 variant. (B) Proportion of dose four effectiveness (against mild disease, hospitalisation, and death) that is attributable to boosting (with either the current or bivalent vaccine product), relative to the proportion of overall efficacy that is attributable to boosting with the bivalent product (rather than the original), for one year following the fourth dose.

## Discussion

As the world transitions towards endemic circulation of SARS-CoV-2, there is a need to continue to evaluate COVID-19 vaccine effectiveness against circulating variants of the virus. Our modelling framework presents a method to integrate the insight that has been generated from understanding the utility of NAT as a correlate of clinical protection with larger population-based cohorts of vaccine effectiveness. By fitting a mechanistic model to such data, it is possible to make short-term projections regarding vaccine effectiveness beyond the period of observation that can help to inform ongoing vaccination strategies and in particular the need for regular boosters for the highest risk populations.

One of the challenges with planning future vaccine booster strategies will be assessing the potential effectiveness of future variant-specific vaccines. Given the speed with which new variants continue to emerge, coupled with the difficulty in identifying appropriate comparator groups to directly estimate vaccine effectiveness, it is likely that decisions will need to be made based on immunogenicity data. For both the Delta and the Omicron variants, applying the fold reductions estimated from immunogenicity data to the relationship inferred against the Wuhan virus in the original paper by Khoury *et al*.^14^ resulted in a more pessimistic prediction of vaccine effectiveness than was inferred in our analysis from fitting to the clinical data. This may be in part due to the uncertainty in the fold reduction given the widespread variation reported across the different laboratory studies. However, it may also indicate that immune responses other than NAT are providing a higher degree of cross-protection against new variants than would be predicted based on NATs alone.

Our results demonstrate that the value of the original vaccines, whilst providing initial high levels of protection, has gradually been diminished through both waning of protection following the 3^rd^ and subsequent doses and the substantial immune escape presented by the Omicron variant. In combination, these two effects combine to generate a substantial additional estimated benefit of switching boosters to the more recent bivalent vaccines – which we estimate to prevent nearly double the number of episodes of severe disease over a 1-year period compared to boosting with the original vaccine. It should be noted that this estimate is sensitive to the underlying shape of our inferred relationship between immune levels and protection, and in particular to the precise point at which there is a rapid drop in protection compared to the “plateau” at high immunity levels. It also depends on the degree to which the new sub-variants that have subsequently emerged exhibit immune escape from the bivalent vaccines. However, early follow-up of the impact of bivalent boosters in the US have demonstrated an added benefit over monovalent boosters.^30^

One of the major limitations of our work is that it was not possible to distinguish the combined effects of infection- and vaccine-induced immunity. The vaccine effectiveness data to which we fit our model is based on the full population of England with effectiveness estimates obtained by comparing outcomes according to vaccine dose against those with no prior vaccination. Given the widespread circulation of the SARS-CoV-2 virus in the community in England throughout the latter half of 2021 and first half of 2022 (from which these data derive), it is likely that a substantial proportion of both the vaccinated and unvaccinated cohorts will have experienced one or more episodes of infection. Thus, these estimates – and our associated short-term projections of vaccine efficacy beyond the observation period – may not hold in other countries with a different background level of infection-induced immunity.

A second limitation in understanding the immune dynamics driving these patterns of vaccine effectiveness was the lack of associated immunological measurements. To overcome this, we inferred immunity levels by treating them as an unobserved process by utilising the parametric forms that have previously been developed to relate NAT to clinical protection. In doing so, we capture the effect of all aspects of the immune response – including both antibody-mediated immunity and potential T-cell responses – in our inferred immunity levels. However, this simple approach, whilst appropriate for short-term parametric projections, may fail to fully capture longer-term immune dynamics. Furthermore, it does not allow us to gain any further mechanistic insight into the underlying immune dynamics driving the observed vaccine effectiveness against the different clinical endpoints. Further research in this area will require careful analysis of large population cohorts containing both immunological and clinical measurements.

One of the challenges with estimating vaccine effectiveness from population cohort data is the difficulty with distinguishing hospitalisations or deaths arising due to COVID-19 from those arising from other causes but in which patients also received a positive COVID-19 diagnosis. The vaccine effectiveness estimates that we used to fit our model are based on hospital admission data with a recent positive diagnosis and hence do not allow us to disentangle this. However, more recent analyses by UKHSA have shown similar vaccine effectiveness estimates when restricting the data to those that were admitted with a respiratory diagnosis which, whilst imperfect, may be more representative of those admitted due to COVID-19.^27^ Nevertheless, this limits our ability to truly assess waning in vaccine effectiveness over time.

Regular booster vaccination is expected to be a key part of the ongoing management of COVID-19 over the coming years, especially among older and more clinically vulnerable populations where protection from any degree of SARS-CoV-2 infection may be crucial. As the SARS-CoV-2 virus continues to evolve, validated models that can predict the effectiveness of modified vaccine products based on immunogenicity data alone will be increasingly important for assessing the benefit of additional doses with either existing or variant-modified vaccines. Our results demonstrate the challenges with doing so, given the more complex immune landscape that has arisen with both past vaccination and ongoing infection with multiple Omicron sub-variants across the world.

## Data Availability

The data used in the fitting were extracted from the cited papers and are provided along with all code for the paper on the Github link.

https://github.com/mrc-ide/covid_efficacy

## Availability of data

The data used in the fitting were extracted from the cited papers and are provided along with all code for the paper at: https://github.com/mrc-ide/covid_efficacy

## Author contributions

Conceptualisation and study design: ACG, ABH, NMF, OJW, GB, PW, SLW; Vaccine efficacy model fitting: NMF, ACG, PD, ABH, DK; Analysis: ACG, ABH, JT, DOM, NMF, PD, SLW; Visualization: ABH, ACG; Writing – original draft: ACG, ABH, NMF, EMR; Writing – review & editing: All authors.

## Supplementary Material

**Table S1:** Estimated vaccine effectiveness against mild disease, hospitalisation and death for Oxford/AstraZeneca AZD1222, Pfizer-BioNTech BNT162b2 and Moderna mRNA-1273 vaccine regimens as a function of time since dose 2 or booster. Estimates are shown for the Delta variant; estimates for the Omicron variant are shown in **Table 2**. Values shown are the posterior median and 95% credible intervals.

| Vaccine | Days post dose 2 |  | Days post booster |  |  |  |  |  |  |
| --- | --- | --- | --- | --- | --- | --- | --- | --- | --- |
|  | 90 | 180 | 30 | 60 | 90 | 120 | 150 | 180 | 365 |
| *Mild disease* |  |  |  |  |  |  |  |  |  |
| Oxford/AstraZeneca AZD1222 | 56.6 (55.6-57.2) | 38.4 (37.2-39.3) | 87.5 (84.2-90.2) | 80.3 (75.4-84.3) | 72.3 (66.2-77.5) | 65.2 (58.3-71.1) | 59.6 (52.4-66) | 55.5 (48.3-62.2) | 44.8 (36.9-52.3) |
| Moderna mRNA-1273 | 82.5 (81.9-83.1) | 69.3 (68.1-70.5) | 94.3 (94.1-94.6) | 90.6 (90.2-91) | 86 (85.5-86.7) | 81.5 (80.8-82.4) | 77.6 (76.7-78.7) | 74.6 (73.5-76) | 65.7 (62-68.6) |
| Pfizer-BioNTech BNT162b2 | 75.8 (75.3-76.3) | 60 (58.8-61) | 92.6 (92.4-92.9) | 87.9 (87.6-88.3) | 82.4 (81.8-83) | 77 (76.2-77.8) | 72.5 (71.5-73.6) | 69.1 (67.8-70.4) | 59.3 (55.2-62.4) |
| *Hospitalisation* |  |  |  |  |  |  |  |  |  |
| Oxford/AstraZeneca AZD1222 | 90.7 (90.2-91.2) | 82.3 (81.2-83.3) | 98.1 (97.5-98.6) | 96.8 (95.9-97.6) | 95.1 (93.7-96.3) | 93.3 (91.4-94.9) | 91.7 (89.3-93.6) | 90.3 (87.6-92.5) | 85.9 (81.6-89.2) |
| Moderna mRNA-1273 | 97.2 (97.1-97.4) | 94.4 (94-94.9) | 99.2 (99.1-99.3) | 98.6 (98.5-98.7) | 97.9 (97.7-98) | 97.1 (96.8-97.3) | 96.3 (96-96.6) | 95.7 (95.3-96.1) | 93.5 (92.4-94.3) |
| Pfizer-BioNTech BNT162b2 | 95.9 (95.7-96.1) | 91.8 (91.2-92.4) | 98.9 (98.9-99) | 98.2 (98.1-98.3) | 97.2 (97.1-97.4) | 96.2 (95.9-96.4) | 95.2 (94.8-95.5) | 94.4 (93.9-94.8) | 91.6 (90.2-92.6) |
| *Death* |  |  |  |  |  |  |  |  |  |
| Oxford/AstraZeneca AZD1222 | 90.4 (89-91.7) | 81.8 (79.4-84) | 98.1 (97.4-98.6) | 96.7 (95.7-97.6) | 95 (93.4-96.3) | 93.1 (91-94.8) | 91.4 (88.8-93.5) | 90 (87-92.5) | 85.4 (80.9-89.1) |
| Moderna mRNA-1273 | 97.1 (96.7-97.6) | 94.2 (93.4-95) | 99.2 (99-99.3) | 98.6 (98.4-98.8) | 97.8 (97.5-98.2) | 97 (96.5-97.4) | 96.2 (95.6-96.8) | 95.5 (94.8-96.2) | 93.3 (91.8-94.4) |
| Pfizer-BioNTech BNT162b2 | 95.8 (95.2-96.4) | 91.5 (90.3-92.7) | 98.9 (98.7-99.1) | 98.1 (97.9-98.4) | 97.1 (96.7-97.6) | 96 (95.4-96.6) | 95 (94.2-95.8) | 94.2 (93.2-95) | 91.3 (89.5-92.8) |

**Figure S1:**
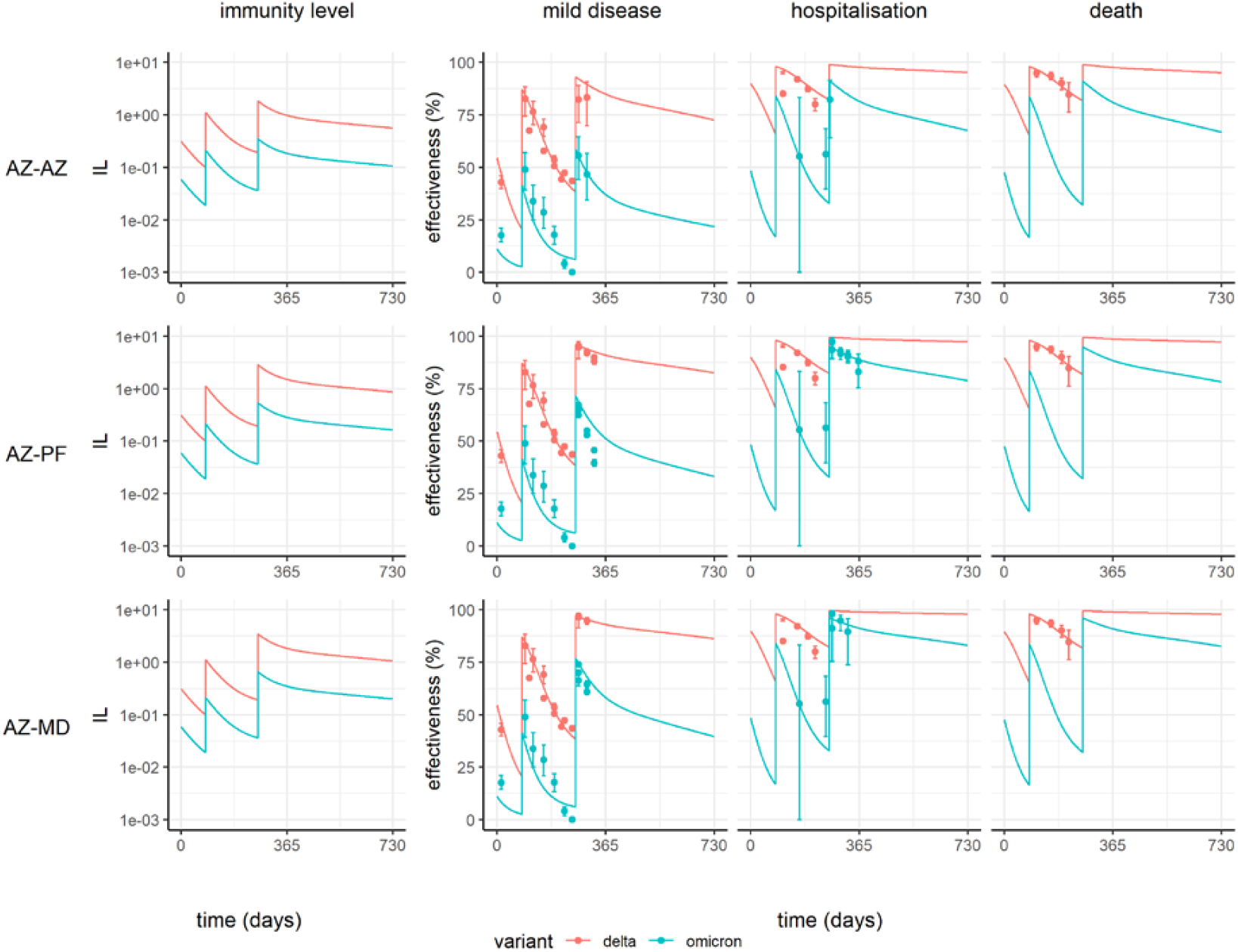
Predicted vaccine effectiveness over time for the Oxford/AstraZeneca AZD1222 (AZ) vaccine delivered as a primary series, followed by boosting with either AZ, or the original Pfizer-BioNTech BNT162b2 (PF) or Moderna mRNA-1273 (MD) vaccine. Plots show neutralizing immunity level (IL) in the left column, alongside effectiveness against mild disease, hospitalisation, and death on the right. Neutralization and protection against the Delta and Omicron variants are shown in red and blue respectively. AZ is delivered for the first two doses, and three booster dose regimens are shown: AZ (AZ-AZ); PF (AZ-PF); and MD (AZ-MD). The solid lines show the posterior median fitted model estimate, and the points show estimates of vaccine effectiveness against three endpoints using data from England.^20,38^

**Table S2:** Statistical fit of the main model and the two sensitivity analyses. Statistics are reported from the sampling phase of the MCMC chain discarding the burn-in period.

| Model | Number of parameters | LogLikelihood (Mean) | LogLikelihood (SE) |
| --- | --- | --- | --- |
| Main | 17 | -79,473 | 3.14 |
| Alternative Severity | 17 | -79,460 | 3.34 |
| Additive Boost | 17 | -79,846 | 3.21 |

**Figure S2:**
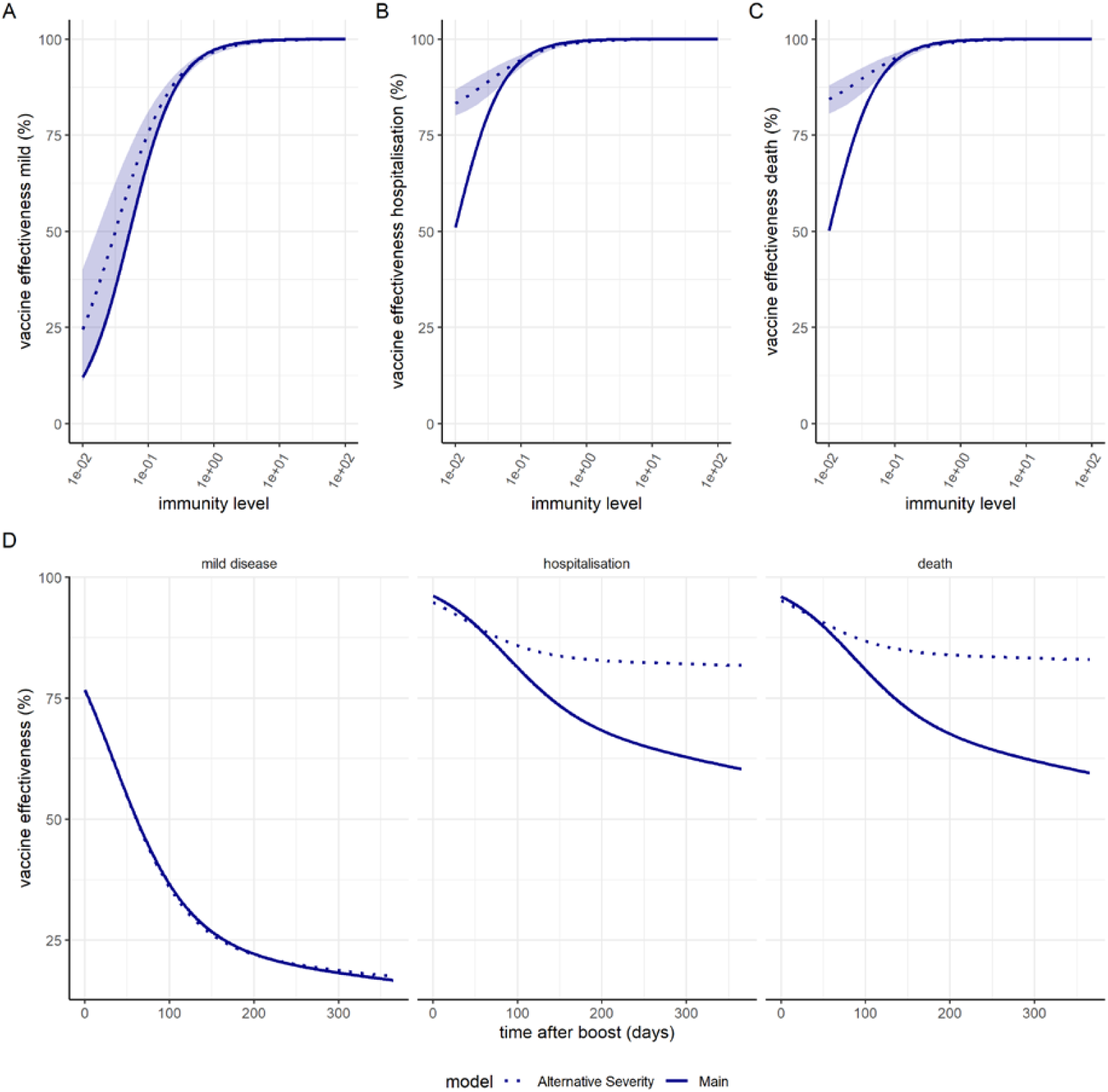
Fitted relationships for the alternative severity model. Dose-response curves estimated from fitting to vaccine effectiveness data for the relationship between immunity level (IL, x-axis) and vaccine effectiveness against mild disease (A), hospitalisation (B) and death (C) against the Omicron/BA.1 variant. The dashed lines show the posterior median estimates and colour bands the 95% credible interval. The solid lines show the dose-response curves using the main model. (D) Predicted vaccine effectiveness for the Moderna mRNA-1273 vaccine from time since dose 3 for the main model (solid line) and the alternative severity model (dashed line). The values are for the posterior median estimates.

**Figure S3:**
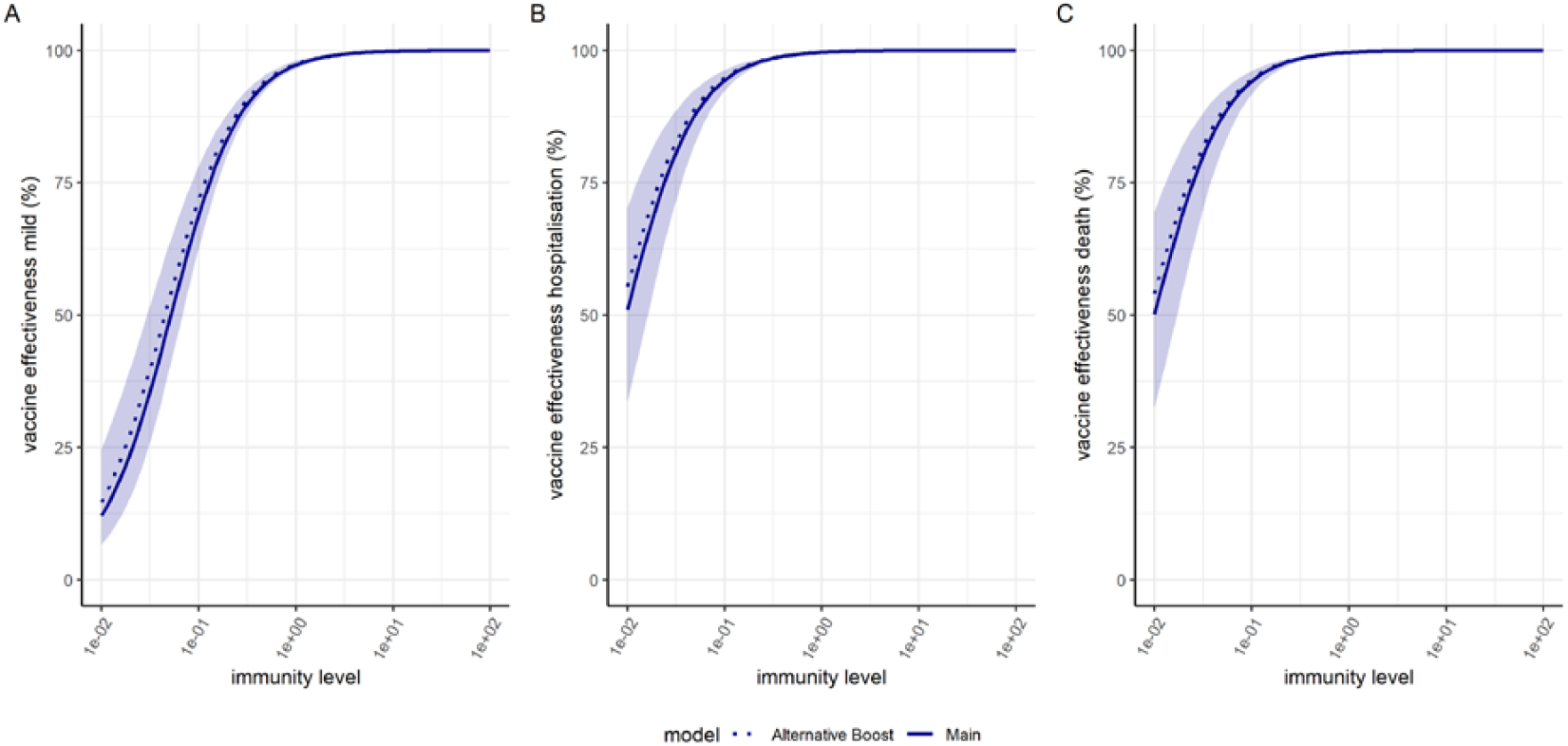
Fitted relationships for the additive boosting model. Dose-response curves estimated from fitting to vaccine effectiveness data for the relationship between immunity level (IL, x-axis) and vaccine effectiveness against mild disease (A), hospitalisation (B) and death (C) against the Omicron/BA.1 variant. The dashed lines show the posterior median estimates and colour bands the 95% credible interval. The solid lines show the dose-response curves using the main model. Given the overlap between the predicted relationships, vaccine effectiveness estimates are similar to those from the main model.

